# Risk of hospitalisation with coronavirus disease 2019 in healthcare workers and their households: a nationwide linkage cohort study

**DOI:** 10.1101/2020.08.03.20164897

**Authors:** ASV Shah, R Wood, C Gribben, D Caldwell, J Bishop, A Weir, S Kennedy, R Martin, A Smith-Palmer, D Goldberg, J McMenamin, C Fischbacher, C Robertson, S Hutchinson, P McKeigue, C Helen, DA McAllister

## Abstract

**Objective:** Many healthcare staff work in high-risk settings for contracting and transmitting Severe Acute Respiratory Syndrome Coronavirus 2. Their risk of hospitalisation for coronavirus disease 2019 (COVID-19), and that of their households, is poorly understood.

**Design and settings and participants:** During the peak period for COVID-19 infection in Scotland (1^st^ March 2020 to 6^th^ June 2020) we conducted a national record linkage study to compare the risk of COVID-19 hospitalisation among healthcare workers (age: 18-65 years), their households and other members of the general population.

**Main outcome:** Hospitalisation with COVID-19

**Results:** The cohort comprised 158,445 healthcare workers, the majority being patient facing (90,733 / 158,445; 57.3%), and 229,905 household members. Of all COVID-19 hospitalisations in the working age population (18-65-year-old), 17.2% (360 / 2,097) were in healthcare workers or their households. Adjusting for age, sex, ethnicity, socio-economic deprivation and comorbidity, the risk of COVID-19 hospitalisation in non-patient facing healthcare workers and their households was similar to the risk in the general population (hazards ratio [HR] 0.81; 95%CI 0.52-1.26 and 0.86; 95%CI 0.49-1.51 respectively). In models adjusting for the same covariates however, patient facing healthcare workers, compared to non-patient facing healthcare workers, were at higher risk (HR 3.30; 95%CI 2.13-5.13); so too were household members of patient facing healthcare workers (HR 1.79; 95%CI 1.10-2.91). On sub-dividing patient-facing healthcare workers into those who worked in front-door, intensive care and non-intensive care aerosol generating settings and other, those in ‘front door’ roles were at higher risk (HR 2.09; 95%CI 1.49-2.94). For most patient facing healthcare workers and their households, the estimated absolute risk of COVID-19 hospitalisation was less than 0.5% but was 1% and above in older men with comorbidity.

**Conclusions:** Healthcare workers and their households contribute a sixth of hospitalised COVID-19 cases. Whilst the absolute risk of hospitalisation was low overall, patient facing healthcare workers and their households had 3- and 2-fold increased risks of COVID-19 hospitalisation.

## Introduction

Severe Acute Respiratory Syndrome Coronavirus 2 (SARS-CoV-2) continues to spread globally, with over 8 million coronavirus disease 2019 (COVID-19) cases and over half a million deaths as of 10^th^ July 2020.^1^

Healthcare workers, who have been integral to the response to COVID-19, may be at increased risk of contracting SARS-CoV-2^2^, and hence subsequently transmitting it to their household and/or workplace contacts.^3^ Estimating the risk in this population is important to guide public health measures to protect healthcare workers and their families, maintain a functioning healthcare system and control rates of secondary transmission within the community.^4^

Despite this, the extent of these risks are not well understood, as the majority of studies have been in single centres,^2^ limited by small sample sizes and/or biased selection and recording of disease.^2 5^ We are well placed to address these limitations in Scotland for two reasons. First, the overwhelming majority of healthcare (especially acute care) is directly delivered by the National Health Service (NHS), which also maintains a national database on all directly employed staff in Scotland including nursing, medical, allied health professionals and support staff. Secondly, Scotland has a well-established health record linkage system.^6–8^

Using record linkage, we evaluated the risk of COVID-19 hospitalisation in healthcare workers in patient and non-patient facing roles along with the risk of their household members. We further evaluated the risk of COVID-19 hospitalisation in patient facing healthcare workers in different clinical settings including intensive care and front-door departments.

## Methods

### Population, data sources and record linkage

Healthcare workers were included if on the 1^st^ of March 2020 (the first positive reported case of COVID-19 in Scotland) they were directly employed by the NHS and/or contracted to provide NHS general practice services in Scotland. Healthcare worker data were extracted from the Scottish Workforce Information Standard System (SWISS) and General Practitioner Contractor Database (GPCD) data **(*Supplementary text 1*)**. Dental and staff working exclusively in paediatric roles were excluded alongside other exclusions due to incomplete or inconsistent data **(*Supplementary text 2*)**. Healthcare worker data were linked to the Community Health Index (CHI) database, a registry of all patients registered to receive care from the NHS in Scotland, close to the complete population. The CHI database includes individuals’ CHI number, a unique patient identifier used on all healthcare records in Scotland.

A cohort was created linking these healthcare worker data to multiple Scottish-wide databases **(*Supplementary figure 1*)** using the CHI number. These included datasets containing individual level clinical information for virology testing for SARS-CoV-2, general hospitalisation data, community prescribing, critical care admissions and the national register for deaths (***Supplementary text 1*)**.

We also used the CHI database to identify all individuals who were not themselves healthcare workers but shared a household with a healthcare worker. Individuals were assigned to the same household if the address (including house and, if included, apartment number) on the CHI database was identical for both; fuzzy matching was not allowed. These household members were then also linked to the Scottish-wide datasets to construct a household member specific cohort **(*Supplementary figure 1*)**. The healthcare worker cohort was restricted to the working-age population (18-65-years-of-age) but the household member cohort included all ages.

Finally, we appended selected variables from the healthcare worker and household member data to an existing Scottish case-control study REACT-COVID-19.^9^ REACT-COVID-19 included linked patient data (excluding healthcare worker and household member status) of **all** cases with a positive SARS-CoV-2 test or COVID-19 as cause of death on certification in Scotland. Each case was matched to 10 age-sex geographically (general practice area) matched controls from the Scottish population A nested case control design was used as this minimises the time required for data processing and computation without loss of statistical power. These data allowed for comparisons with the general population – defined as residents of Scotland who were not healthcare workers or members of their households.

### Outcomes

Outcomes were restricted to the time period from the 1^st^ of March to the 6^th^ of June 2020. The primary outcome was COVID-19 hospitalisation defined as the first positive test for SARS-CoV-2 in hospital and/or the individual being hospitalized within 28 days of testing positive. Secondary outcomes reported were admission to intensive care and death occurring within 28 days of first testing positive. Tests were included whether they were undertaken for screening or clinical purposes. Hospitalisation was chosen as the primary outcome because milder disease not requiring hospitalisation is likely to suffer from ascertainment bias (since healthcare workers may be more likely to be tested), and because hospitalisation with COVID-19 is a clinically significant event.

### Exposure

Occupational roles were defined for all healthcare workers using the SWISS/GPCD databases. Broad roles were categorised into either patient facing, non-patient facing or undetermined. Roles were defined based on formal job titles for nursing, allied health professionals and support staff and specialty for medical staff. Selected nursing staff were additionally assigned on the basis of their working location (for example the emergency department). These definitions were deliberately narrow, assigning around a fifth of healthcare workers to “undetermined” **(*Supplementary text 3*)**. This was done to avoid non-differential misclassification bias.

Patient facing roles were further divided into the following settings; ‘front-door’ (for example paramedics or workers in acute receiving specialties), ‘intensive care’ (ICU), non-intensive care but still exposed to aerosol generating procedures (AGP) (for example workers in respiratory medicine) and ‘other’. These designations were made prior to database linkage (see statistical analysis plan). Household members were assigned to the role of the associated member of staff. Where there was more than more than one healthcare worker in a household, the highest risk designation was applied.

### Covariates

Occupation-related covariates obtained from the healthcare worker database were seniority grade, occupation (medical, nursing, allied health professional, support, administration and other), length of service, immigration status, and full/part-time working status. Age, sex and the Scottish Index of Multiple Deprivation (SIMD) quintile, an area-based measure of socio-economic deprivation^10^ were obtained from the CHI register. Comorbidities were identified using predefined criteria from previous hospitalisations **(*Supplementary text 4*)** and/or recently dispensed medications. Ethnicity was recorded across multiple datasets defined using the ONOMAP algorithm.^11^

### Statistical analysis

The cumulative incidence of COVID-19 hospitalisation was plotted for healthcare workers, household members and working-age adults in the general population who were not healthcare workers or their household members. The denominator for the latter was obtained by subtracting the healthcare worker and household cohorts from the 2019 mid-year estimates. In the healthcare worker and household cohorts we modelled COVID-19 hospitalisation using Cox regression, calculating robust standard errors to allow for clustering due to shared household membership and stratifying on groups of health board areas to allow for differences in baseline hazard. These strata were chosen a priori based on data for the general Scottish population.

In the case-control study we performed conditional logistic regression. Since REACT-COVID-19^9^ used incidence density sampling, the effect measure estimates derived from these case-control analysis are directly comparable to those derived from the Cox regression. To allow comparison against the general population across the cohort and case-control analyses, the non-patient facing role was used as a common reference group.

A separate prespecified statistical analysis plan has also been provided. Analyses were conducted in R version 3.6.1 (Vienna, Austria). Analysis code will be made available here – [WILL BE MADE PUBLIC PRIOR TO MANUSCRIPT PUBLICATION].

### Patient and Public Involvement

This project was approved by the Public Benefit and Privacy Panel (2021-0013). Further, both the British Medical Association and the Royal College of General Practitioners in Scotland – who in this case were representatives of some of the subjects of this research – provided a letter in support of the project.

## Results

There were 158,445 healthcare workers and 229,905 household members. The majority of healthcare workers (124,661/158,445; 78.7%), but only 88,274 (38.4%) of household members, were women. Over half of healthcare workers (90,733/158,445; 57.3%) were patient-facing with 32,615 (20.6%) and 35,097 (22.2%) classified as non-patient facing and undetermined respectively **(*Table 1*)**. Most patient facing healthcare workers were in front door roles **(*Supplementary table 1*)**.

**Table 1:** Baseline characteristics of healthcare workers and members of their households

|  | Healthcare workers |  |  |  | Household members of healthcare workers |  |  |  |
| --- | --- | --- | --- | --- | --- | --- | --- | --- |
|  | Total | Non patient-facing healthcare workers | Undetermined healthcare workers | Patient facing healthcare workers | Total household members of healthcare workers | Household of non-patient facing healthcare workers | Household of undetermined healthcare workers | Household of patient facing healthcare workers |
| Number | 158445 | 32615 | 35097 | 90733 | 229905 | 44812 | 48530 | 136563 |
| Age, mean(sd) | 44.49 (11.56) | 46.64 (11.09) | 45.7 (11.7) | 43.24 (11.51) | 30.88 (20.93) | 33.19 (21.19) | 31.86 (20.98) | 29.77 (20.74) |
| Sex n(%) |  |  |  |  |  |  |  |  |
| Female | 124661 (78.7%) | 26299 (80.6%) | 25916 (73.8%) | 72446 (79.8%) | 88274 (38.4%) | 16589 (37%) | 18651 (38.4%) | 53034 (38.8%) |
| SIMD quintile n(%)* |  |  |  |  |  |  |  |  |
| 1 (least deprived) | 24066 (15.2%) | 4895 (15%) | 6773 (19.3%) | 12398 (13.7%) | 31337 (13.6%) | 5935 (13.2%) | 8510 (17.5%) | 16892 (12.4%) |
| 2 | 29894 (18.9%) | 6338 (19.4%) | 7100 (20.2%) | 16456 (18.1%) | 41466 (18%) | 8457 (18.9%) | 9533 (19.6%) | 23476 (17.2%) |
| 3 | 31213 (19.7%) | 6465 (19.8%) | 6570 (18.7%) | 18178 (20%) | 44291 (19.3%) | 8621 (19.2%) | 8735 (18%) | 26935 (19.7%) |
| 4 | 35528 (22.4%) | 7431 (22.8%) | 6970 (19.9%) | 21127 (23.3%) | 53197 (23.1%) | 10456 (23.3%) | 10117 (20.8%) | 32624 (23.9%) |
| 5 (most deprived) | 37744 (23.8%) | 7486 (23%) | 7684 (21.9%) | 22574 (24.9%) | 59614 (25.9%) | 11343 (25.3%) | 11635 (24%) | 36636 (26.8%) |
|  | Total | Non patient-facing healthcare workers | Undetermined healthcare workers | Patient facing healthcare workers | Total household members of healthcare workers | Household of non-patient facing healthcare workers | Household of undetermined healthcare workers | Household of patient facing healthcare workers |
| **Ethnic group, n(%)** |  |  |  |  |  |  |  |  |
| White | 153126 (96.6%) | 31991 (98.1%) | 33813 (96.3%) | 87322 (96.2%) | 219914 (95.7%) | 43445 (96.9%) | 46163 (95.1%) | 130306 (95.4%) |
| South Asian | 3704 (2.3%) | 453 (1.4%) | 899 (2.6%) | 2352 (2.6%) | 6600 (2.9%) | 919 (2.1%) | 1594 (3.3%) | 4087 (3%) |
| Black | 657 (0.4%) | 70 (0.2%) | 150 (0.4%) | 437 (0.5%) | 1385 (0.6%) | 157 (0.4%) | 309 (0.6%) | 919 (0.7%) |
| Chinese | 462 (0.3%) | 40 (0.1%) | 115 (0.3%) | 307 (0.3%) | 672 (0.3%) | 89 (0.2%) | 170 (0.4%) | 413 (0.3%) |
| Other | 496 (0.3%) | 61 (0.2%) | 120 (0.3%) | 315 (0.3%) | 1334 (0.6%) | 202 (0.5%) | 294 (0.6%) | 838 (0.6%) |
| **Comorbidity, n(%)** | 21143 (13.3%) | 4701 (14.4%) | 4864 (13.9%) | 11578 (12.8%) | 20978 (9.1%) | 4599 (10.3%) | 4867 (10%) | 11512 (8.4%) |
| Ischaemic heart disease | 1614 (1%) | 338 (1%) | 461 (1.3%) | 815 (0.9%) | 2798 (13.3%) | 634 (13.8%) | 676 (13.9%) | 1488 (12.9%) |
| Other heart disease | 3726 (2.4%) | 812 (2.5%) | 854 (2.4%) | 2060 (2.3%) | 4577 (21.8%) | 1037 (22.5%) | 1063 (21.8%) | 2477 (21.5%) |
| Other circulatory system diseases | 2450 (1.5%) | 487 (1.5%) | 593 (1.7%) | 1370 (1.5%) | 2967 (14.1%) | 644 (14%) | 721 (14.8%) | 1602 (13.9%) |

|  | **Healthcare workers** |  |  |  | **Household members of healthcare workers** |  |  |  |
| --- | --- | --- | --- | --- | --- | --- | --- | --- |
|  | **Total** | **Non patient-facing healthcare workers** | **Undetermined healthcare workers** | **Patient facing healthcare workers** | **Total household members of healthcare workers** | **Household of non-patient facing healthcare workers** | **Household of undetermined healthcare workers** | **Household of patient facing healthcare workers** |
| Advanced chronic kidney disease | 110 (0.1%) | 41 (0.1%) | 19 (0.1%) | 50 (0.1%) | 160 (0.8%) | 26 (0.6%) | 42 (0.9%) | 92 (0.8%) |
| Asthma and chronic lower respiratory disease | 3349 (2.1%) | 705 (2.2%) | 771 (2.2%) | 1873 (2.1%) | 4169 (19.9%) | 861 (18.7%) | 965 (19.8%) | 2343 (20.4%) |
| Neurological disorders | 691 (0.4%) | 173 (0.5%) | 129 (0.4%) | 389 (0.4%) | 1011 (4.8%) | 216 (4.7%) | 229 (4.7%) | 566 (4.9%) |
| Severe liver disease | 96 (0.1%) | 15 (0%) | 27 (0.1%) | 54 (0.1%) | 134 (0.6%) | 29 (0.6%) | 29 (0.6%) | 76 (0.7%) |
| Malignant Neoplasms | 5662 (3.6%) | 1323 (4.1%) | 1269 (3.6%) | 3070 (3.4%) | 3880 (18.5%) | 853 (18.5%) | 898 (18.5%) | 2129 (18.5%) |
| Disorders of esophagus, stomach and duodenum | 3886 (2.5%) | 872 (2.7%) | 854 (2.4%) | 2160 (2.4%) | 3496 (16.7%) | 776 (16.9%) | 787 (16.2%) | 1933 (16.8%) |
| Diabetes, type 1 | 1080 (0.7%) | 249 (0.8%) | 227 (0.6%) | 604 (0.7%) | 1204 (5.7%) | 250 (5.4%) | 234 (4.8%) | 720 (6.3%) |
| Diabetes, type 2 | 3763 (2.4%) | 944 (2.9%) | 972 (2.8%) | 1847 (2%) | 4762 (22.7%) | 1093 (23.8%) | 1210 (24.9%) | 2459 (21.4%) |
| Diabetes (type unknown) | 284 (0.2%) | 80 (0.2%) | 58 (0.2%) | 146 (0.2%) | 302 (0.1%) | 72 (0.2%) | 72 (0.1%) | 158 (0.1%) |
|  | **Total** | **Non patient-facing healthcare workers** | **Undetermined healthcare workers** | **Patient facing healthcare workers** | **Total household members of healthcare workers** | **Household of non-patient facing healthcare workers** | **Household of undetermined healthcare workers** | **Household of patient facing healthcare workers** |
| **Comorbidity count, n(%)** |  |  |  |  |  |  |  |  |
| 0 | 137302 (86.7%) | 27914 (85.6%) | 30233 (86.1%) | 79155 (87.2%) | 208927 (90.9%) | 40213 (89.7%) | 43663 (90%) | 125051 (91.6%) |
| 1 | 16924 (10.7%) | 3714 (11.4%) | 3858 (11%) | 9352 (10.3%) | 31337 (13.6%) | 5935 (13.2%) | 8510 (17.5%) | 16892 (12.4%) |
| >= 2 | 4219 (2.7%) | 1006 (2.9%) | 987 (3%) | 2226 (2.5%) | 15465 (6.7%) | 3395 (7.6%) | 3526 (7.3%) | 8544 (6.3%) |
| **Immigration Status, n(%)** |  |  |  |  |  |  |  |  |
| UK National | 152637 (96.3%) | 34955 (99.6%) | 32550 (99.8%) | 85132 (93.8%) | - | - | - | - |
| Non UK National | 804 (0.5%) | 142 (0.4%) | 65 (0.2%) | 597 (0.7%) | - | - | - | - |
| **Whole or Part Time, n(%)\*** |  |  |  |  | - | - | - | - |
| Whole time | 88634 (55.9%) | 17232 (49.1%) | 20221 (62%) | 51181 (56.4%) | - | - | - | - |
| Part time | 64807 (40.9%) | 17865 (50.9%) | 12394 (38%) | 34548 (38.1%) | - | - | - | - |

Abbreviation: SIMD – Scottish Index of Multiple Deprivation. \*Immigration status and whole or part time status are not included in the GPCD database hence the percentages do not sum to 100% for these variables.

The total Scottish population was estimated at 5,463,300 with the working age population (18-65-years-of-age) estimated at 3,452,592 **(*Supplementary figure 2*)**. Across the entire Scottish population there were a total of 6,346 hospitalisations with COVID-19 **(*Table 2 and Supplementary Figure 2*)**. REACT-COVID-19^9^ included clinical data on **all** these cases and ten randomly selected controls **(*Supplementary Figure 2*)**. Of 6,346 hospitalisations with COVID-19 in Scotland, 33% (n=2,097), occurred in the working age-population (aged 18-65-years). Of these 1,737 (82.8%) occurred in the general population, while healthcare workers and their household members accounted for 243 (11.6%) and 117 (5.6%) respectively **(*Table 2 and Table 3*)**. This meant that healthcare workers and their household members accounted for 17.2% (360/2,097) of COVID-19 hospitalisations while representing only 11.2% (388,350 /3,452,592) of the working age population. Within household members there were a further 24 hospitalisations in 89,327 individuals below the age of 18 or above 65 years **(*Table 3*)**.

**Table 2:** Association between role and risk of COVID 19 hospitalization among healthcare workers, members of their households and the population of Scotland

|  | Healthcare worker comparison |  |  |  | Household member of healthcare worker comparison |  |  |  |
| --- | --- | --- | --- | --- | --- | --- | --- | --- |
|  | General population - other working age residents of Scotland | Non-patient facing | Undetermined | Patient facing | General population - other residents of Scotland (all ages) | Household of non-patient facing healthcare workers | Household of undetermined healthcare workers | Household of patient facing healthcare workers |
| Hospitalized (n) | 1737 | 23 | 39 | 181 | 5962 | 20 | 32 | 89 |
| Total population (n) | 3,153,569 | 32615 | 35097 | 90733 | 5074950 | 44812 | 48530 | 136563 |
| Risk of hospitalization (%) | 0.06 | 0.07 | 0.11 | 0.20 | 0.12 | 0.04 | 0.07 | 0.07 |
| Model 1, Age and sex | 0.92 (0.59-1.42) | 1 | 1.64 (0.99-2.72) | 3.31 (2.13-5.13) | 0.87 (0.50-1.49) | 1 | 1.68 (0.95-2.95) | 1.82 (1.12-2.96) |
| Model 2, as model 1 plus socioeconomic deprivation and ethnicity | 0.90 (0.58-1.40) | 1 | 1.54 (0.93-2.57) | 3.29 (2.12-5.10) | 0.83 (0.48-1.43) | 1 | 1.63 (0.93-2.87) | 1.80 (1.11-2.93) |
| **Model 3 as model 2, plus comorbidity** | **0.81 (0.52-1.26)** | **1** | **1.55 (0.93-2.59)** | **3.30 (2.13-5.13)** | **0.86 (0.49-1.51)** | **1** | **1.60 (0.91-2.82)** | **1.79 (1.10-2.91)** |
| Model 4, as model 3 plus occupation and grade | - | 1 | 1.53 (0.91-2.56) | 3.00 (1.68-5.36) | - | 1 | 1.13 (0.36-3.53) | 1.60 (0.53-4.82) |
| Model 5, as model 4 plus part time status | - | 1 | 1.60 (0.96-2.69) | 3.06 (1.73-5.43) | - | 1 | 1.19 (0.39-3.66) | 1.60 (0.53-4.76) |

Estimates and 95% confidence intervals were obtained from Cox models for the within-healthcare worker and household member comparisons and from conditional logistic regression models for the population versus non patient facing comparison. The conditional logistic regression models were fit within an incidence-density case control study. As such these estimates can be interpreted as hazard ratios. The Cox models were fitted on the entire cohort of healthcare workers and household members respectively. We generated robust standard errors in the Cox models to allow for correlations due to multi-person households. The population denominators were obtained by subtracting the 2019 mid-year population estimates from the number of healthcare workers and their household members.

**Table 3:** Characteristics of cases hospitalized with COVID-19 among healthcare workers, members of their households and the working age population of Scotland. *Cells with count less than 5 will appear as <5 in accordance with disclosure guidance

|  | **Population**<br>**(working age)** | **Healthcare workers\*** | **Household**<br>**members of**<br>**healthcare**<br>**worker\*** |
| --- | --- | --- | --- |
| **Number** | **1737** | **243** | **141** |
| **Age (mean, sd)** | **52.5 (10.5)** | **49.2 (10.1)** | **53.9 (15.0)** |
| **Age strata, n(%)** |  |  |  |
| <18 | - | - | 5 (3.5%) |
| 18-65 | 1737 (100.0%) | 243 (100.0%) | 117 (83.6%) |
| >65 | - | - | 19 (13.6%) |
| **Male (%)** | **953 (54.9%)** | **75 (30.9%)** | **113 (80.7%)** |
| **Comorbidity, n(%)** |  |  |  |
| Ischaemic heart disease | 44 (2.6%) | 8 (3.3%) | 6 (4.3%) |
| Other heart disease | 12 (0.7%) | <5 | <5 |
| Other circulatory system diseases | 3 (0.2%) | 0 | 0 |
| Asthma and chronic lower respiratory disease | 7 (0.4%) | <5 | 0 |
| Neurological disorders | 4 (0.2%) | 0 | 0 |
| Malignant neoplasms | 6 (0.3%) | 0 | 0 |
| Disorders of esophagus, stomach and duodenum | 1 (0.1%) | 0 | 0 |
| Diabetes, type 1 | 8 (0.5%) | <5 | <5 |
| Diabetes, type 2 | 150 (8.7%) | 15 (6.2%) | 18 (12.9%) |
| Diabetes, type unknown | 5 (0.3%) | <5 | <5 |
| **Any comorbidity (%)** | **219 (12.7%)** | **28 (11.5%)** | **27 (19.3%)** |
| **Critical care admission or death, n(%)** |  |  |  |
| Intensive care (%) | 279 (16.1%) | 30 (12.3%) | 28 (20.0%) |
| Died (%) | 227 (13.1%) | 6 (2.5%) | 18 (12.9%) |

### Risk of COVID-19 hospitalisation in healthcare workers

The risk of COVID-19 hospitalisation was 0.20% (181/90,733), 0.07% (23/32,615) and 0.11% (39/35,097) in patient facing, non-patient facing and undetermined healthcare workers respectively **(*Figure 1a*)**.

**Figure 1.**
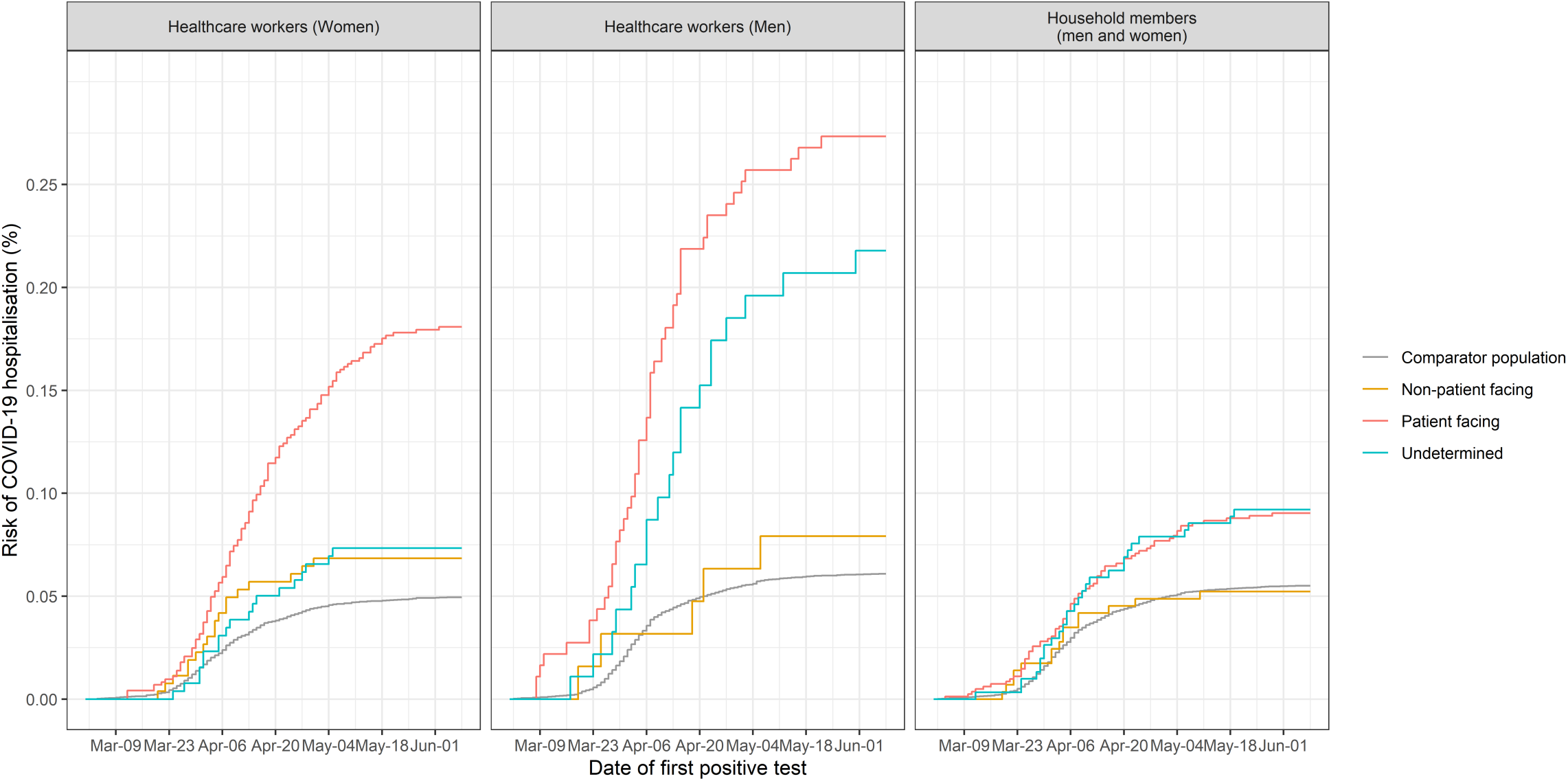
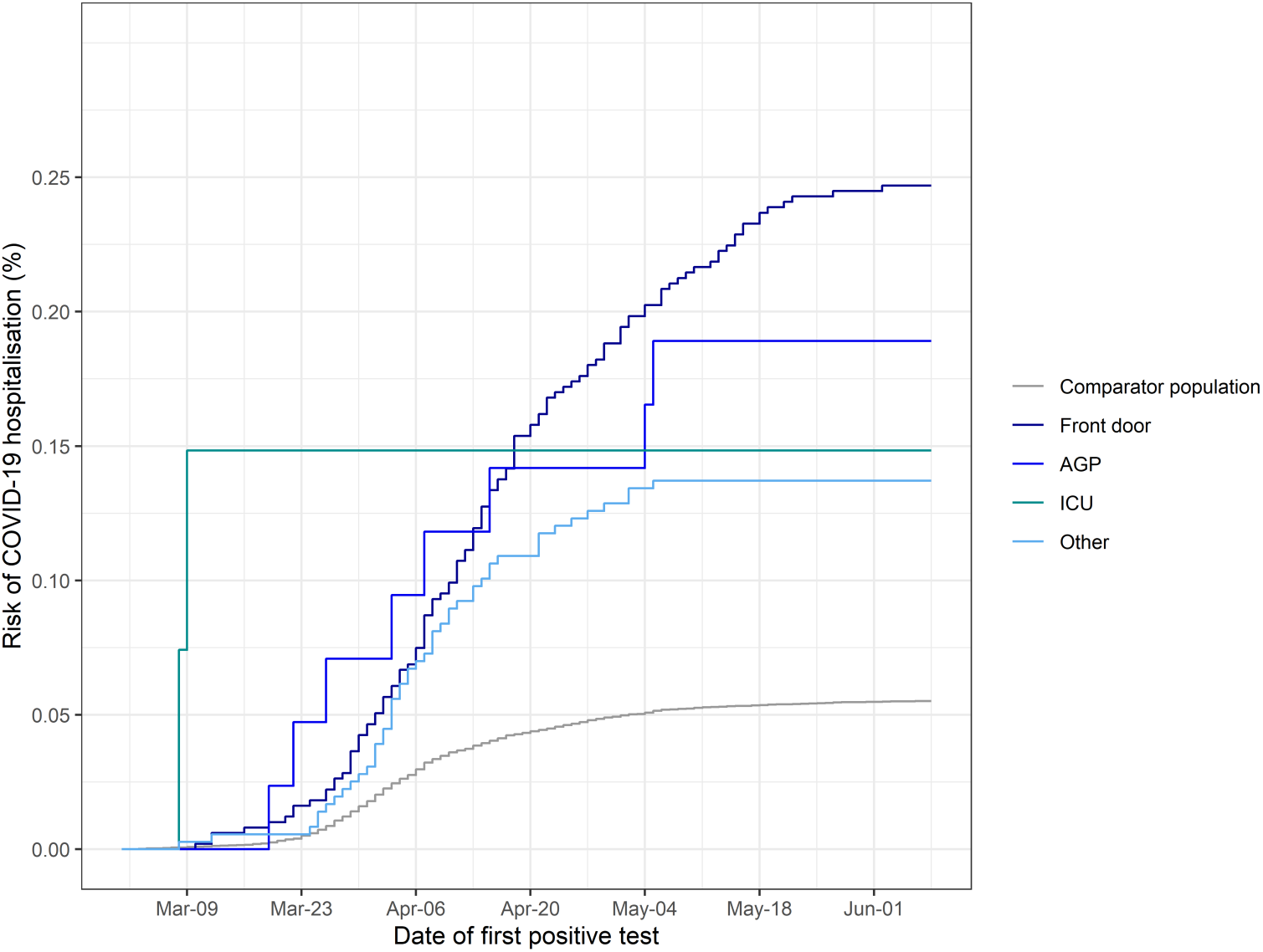
a: Cumulative incidence (risk) of hospitalizations with COVID-19 in healthcare workers, household members of healthcare workers and the general working-age population b: Cumulative incidence (risk) of hospitalizations with COVID-19 in patient facing healthcare workers by specific role

Compared to non-patient facing healthcare workers, after adjusting for age, sex, socio-economic status, ethnicity and comorbidity, patient facing workers were at a higher risk of hospitalisation (hazard ratio [HR] 3.30; 95% confidence interval [CI] 2.13-5.13, ***Table 2, Supplementary Table 2***). There was no evidence of interaction (on the relative scale) by age, sex or comorbidity (p-values 0.57, 0.15 and 0.55 respectively). Adjusting for age, sex, socio-economic deprivation and comorbidity, within healthcare workers in patient-facing roles, compared to those in the ‘other’ category, ‘front door’ workers were more likely to be hospitalized (HR 2.09; 95%CI 1.49-2.94). For workers in (non-intensive care) AGP roles the risk was similarly increased, although the confidence interval included the null (HR 1.91; 95%CI 0.90-4.07). Only 1,348 healthcare workers were assigned to the ‘intensive care’ category, among whom there were less than five hospitalisations all occurring at an early stage of the pandemic (HR 1.22; 95%CI 0.29-5.09, ***Figure 1b***).

Compared to the general population, the risk among non-patient facing healthcare workers was not increased, including after adjusting for age, sex, socio-economic deprivation and comorbidity (HR 0.81; 95%CI 0.52-1.26 ***Table 2***). Healthcare workers with an ‘undetermined’ role had an intermediate level of risk between that of patient facing and non-patient-facing healthcare workers.

In the cumulative incidence plots **(*Figure 1a*)**, the risk in patient-facing workers appeared to plateau earlier in non-patient facing healthcare workers and in the general population than in patient facing healthcare workers. In exploratory analyses we therefore compared the risk in patient facing healthcare workers to the general population (there were too few cases in May for models comparing non-patient facing healthcare workers to converge) over time; conditioning on age and sex the hazard ratios were 2.64; 95%CI (1.82-3.82), 4.18; 95%CI (3.29-5.30) and 6.44; 95%CI (4.00-10.37) for March, April and May respectively (p-value for interaction 0.01).

### Risk of COVID-19 hospitalisation in household members of healthcare workers

The risk of COVID-19 hospitalisation was 0.07% (89/136,563), 0.04% (20/44,812) and 0.07% (32/48,530) in household members of patient facing, non-patient facing and undetermined healthcare workers respectively **(*Figure 1a*)**. The overall absolute risk in household members of healthcare workers below the age of 18 years was low (5/78,253, 0.01%).

Associations seen among household members were similar, albeit attenuated, to those seen among healthcare workers. In models adjusting for age and sex, compared to household members of non-patient facing healthcare workers, those in households of patient facing healthcare workers had a higher risk of hospitalisation (HR 1.82; 95%CI 1.12-2.96). This association was also seen after adjusting for age, sex, ethnicity, socio-economic deprivation and comorbidity (HR 1.79; 95%CI 1.10-2.91). Those in households of non-patient facing healthcare workers had a similar risk to that seen in the general population (0.86; 95% CI 0.49-1.51, ***Table 2, Supplementary table 3***).

### Age, sex and comorbidity

***Figure 2*** illustrates the absolute 90-day risk (from the 1st of March 2020) to healthcare workers and their household members based on Cox models adjusting for role, age, sex and comorbidity. For the majority of healthcare workers and household members the risks remained below 0.5%. Only older men with at least one comorbidity who were in patient facing roles, or who were household member of a patient facing healthcare worker, had risks approaching 1% or higher. Among patient facing healthcare workers, 5% (4,614/90,733) had a household member, or were themselves, in this higher-risk group (male, aged 60 years with one or more comorbidity).

**Figure 2.**
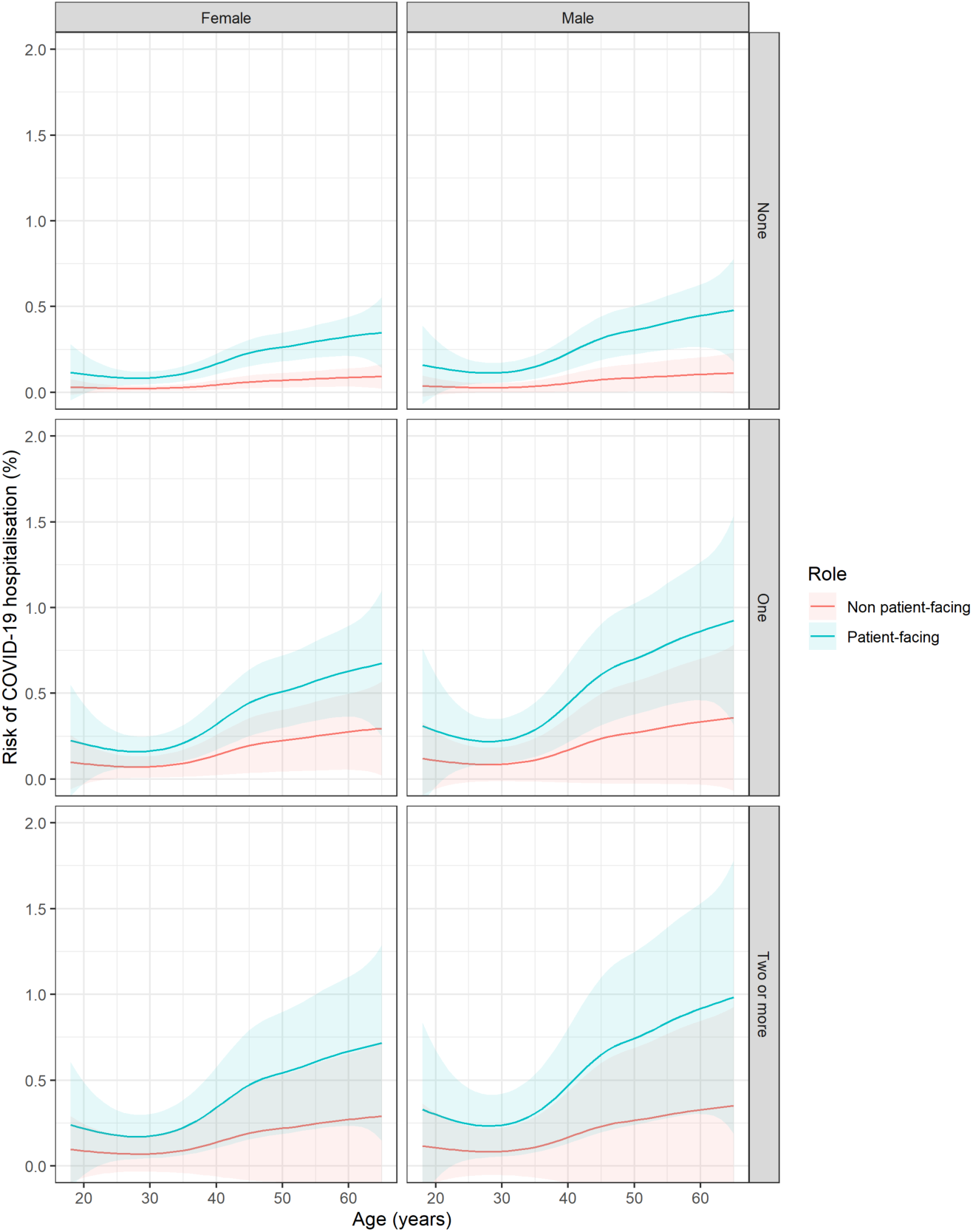
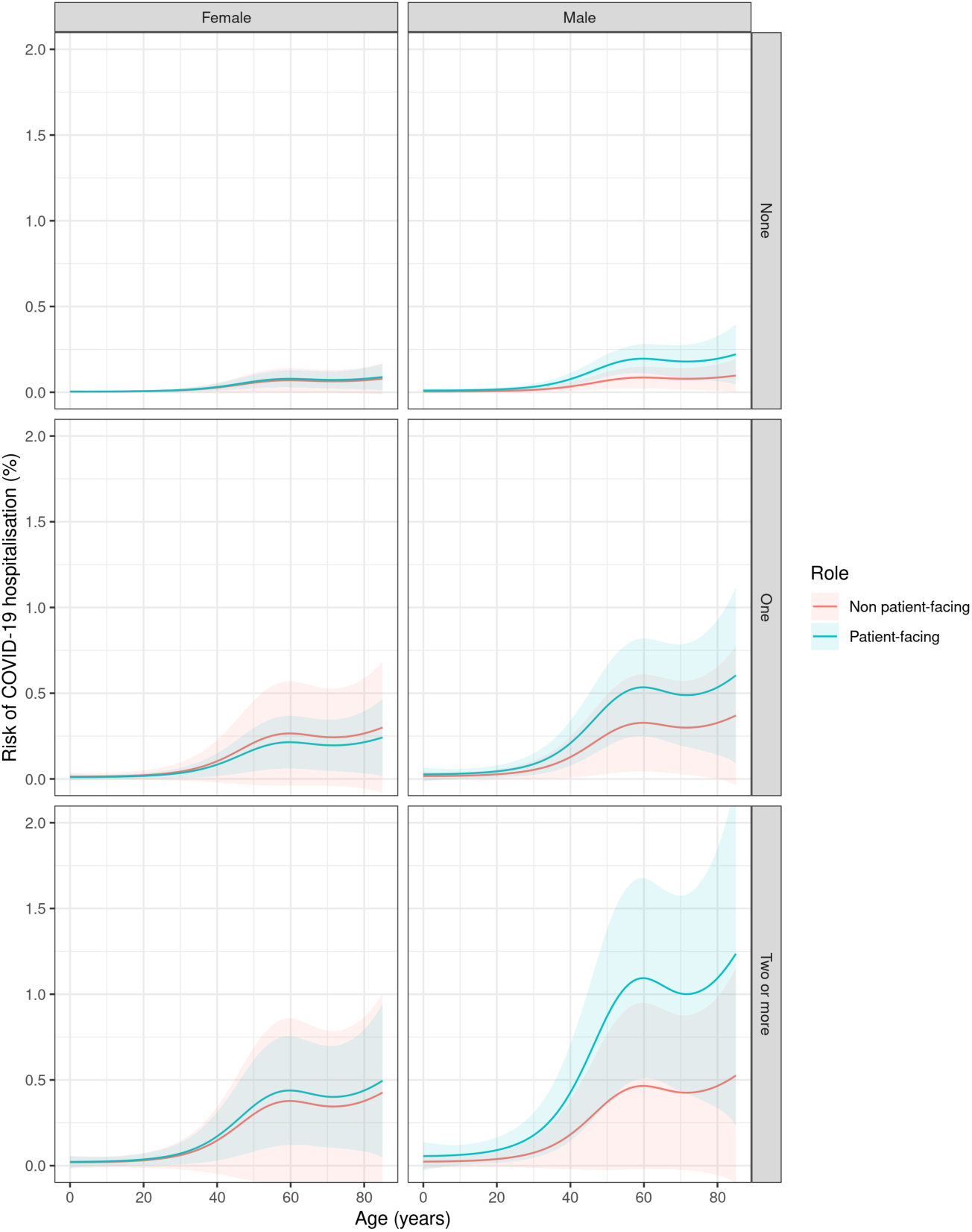
a: 90-day risk of COVID 19 hospitalization from 1st March 2020 by age, sex comorbidity count (none, one or two or more), and occupational role in healthcare workers. Central estimates and 95% confidence intervals were obtained from Cox regression models on age (with penalised splines to allow for non-linearity), sex and comorbidity count. b: 90-day risk of COVID 19 hospitalization from 1st March 2020 by age, sex comorbidity count (none, one or two or more), and occupational role in household members of healthcare workers. Central estimates and 95% confidence intervals were obtained from Cox regression models on age (with penalised splines to allow for non-linearity), sex and comorbidity count.

### Characteristics and outcomes of hospitalised COVID-19 cases in healthcare workers, household members and the general population

Among COVID-19 hospitalisations, compared to the general population, healthcare workers were similar in terms of age and comorbidity (***Table 3***). There were however lower rates of admission to intensive care (30 (12.3%) in healthcare workers and 279 (16.1%) in the working age population) and a lower proportion of deaths occurring within 28 days (6 (2.5%) versus 227 (13.1%)). Household members were more similar to the general population.

## Discussion

In nearly 160,000 healthcare workers and 250,000 household members of healthcare workers we found that hospitalisation with COVID-19 was uncommon, with overall risks of less than half of one percent during the COVID-19 pandemic period (1^st^ of March 2020 to 6^th^ June 2020). Compared to other working age adults, however, this risk was higher. Accounting for age, sex and other confounders, patient facing healthcare workers and members of their households were, respectively, 3-fold and 2-fold more likely to be hospitalized. Healthcare workers and their households accounted for one in six of all COVID-19 hospitalisations in the working age population (18-65 years).

A non-trivial proportion of these hospitalisations resulted in critical care admission or death. Among hospitalized healthcare workers, 1 in 8 were admitted into critical care and 6 (2.5%) died; in hospitalized household members 1 in 5 were admitted to critical care and 18 (12.9%) died. Therefore, as well as having implications for the transmission of COVID-19^3 12^, and the sustainability and deliverability of healthcare^4^, these findings have implications for the safety and well-being of healthcare workers, and their households.^13^

To our knowledge we are the first to report the risk of COVID-19 in nearly 250,000 household members of healthcare workers. Previous evidence on COVID-19 on the risk to household members of healthcare workers is sparse,^14^ despite evidence that their safety is of major importance to healthcare workers.^13^ We show that the risk of COVID-19 hospitalisation was nearly 2-fold higher in household members of patient compared to non-patient facing healthcare workers. Therefore, the susceptibility of household members, as well as healthcare workers themselves need to be considered when assessing occupational risk.

Several studies have reported an increased risk of COVID-19 infection and high prevalence of SARS-CoV-2 in healthcare workers, especially in front-line workers.^2 5 14–16^ However, many of these reports have been were small, single centre, cross-sectional in nature, used methods highly susceptible to bias or restricted their populations to physicians and nurses.^2 5 17 18^ In a large healthcare worker population including a wide range of occupations with robust adjustment for confounding factors, we provide strong evidence that patient facing healthcare workers are at moderately increased risk of experiencing a sufficiently severe form of COVID-19 to require hospitalisation. We provide further evidence that within patient facing healthcare workers, those categorised as working within ‘front door’ specialties, are at the highest risk of COVID-19 hospitalisation, likely reflecting the higher seroprevalence rates of SARS-CoV-2 in this population.^19^

In response to emerging evidence and international guidance, the NHS in Scotland introduced several changes to infection prevention and control guidance during the course of the pandemic.^20^ Despite this, the differential in risk between the general working age population (who had at this time minimal contacts outside their own households) and patient-facing healthcare workers did not fall and may have increased. In contrast, the risk did appear to fall quickly in the “higher risk” intensive care settings. Consistent with international guidance, the NHS in Scotland recommends higher levels of personal protective equipment in higher risk settings, such as intensive care.^22^ In this context, it is notable that less than five healthcare workers based in intensive care were hospitalised, all of whom first tested positive for SARS-CoV-2 in early March. In view of the small numbers of staff in intensive care settings, considerable caution is needed in interpreting this finding, but it is consistent with a recent report from Wuhan that no healthcare workers in high-risk clinical areas tested positive for SARS-CoV-2 in the context of strong infection control measures.^21^ Together with the observations that the relative risk, compared to the general population, in patient facing healthcare workers continued to rise during the course of pandemic, and that the overall risk was highest in front-door healthcare workers, these findings raise particular concerns about moderate exposure settings – both in terms of the risk to staff, and the risk of transmitting infection to the wider community.

In moderate-risk settings, where patients may only have suspected, or even unsuspected COVID-19, the use of more resource-intensive and burdensome personal protective equipment of the kind deployed in high-risk settings is very challenging.^22 23^ One proposed alternative, or additional, measure to improve safety is therefore to redeploy healthcare workers from patient facing to non-patient facing roles if they or their households are more susceptible to severe disease. Our findings suggest that this may be a feasible policy for two reasons. First, non-patient facing healthcare workers and their households had similar risks of hospitalisation as did the general population. Secondly, the proportion of patient facing healthcare workers who themselves, or whose households, were at increased risk of hospitalisation (up to 1%) was low, being around 1 in 20.

Given the small number of deaths in the healthcare worker population we were unable to estimate the risk of COVID-19 related mortality compared to the general population. The Office of National Statistics in England did not find an increased COVID-19 mortality among healthcare workers.^24^ There are several reasons why hospitalisation might be increased without an increase in deaths. While we identified a cohort of healthcare workers, and sub-divided these by occupational roles, finding a risk only in patient-facing healthcare workers, the ONS study relied on self-reporting for the population at risk with information provided by the next of kin at registration. The Office of National Statistics also reported mortality for healthcare workers regardless of their role.^24^ Further, healthcare workers may present earlier improving their survival for a given severity of COVID-19, and/or they may have a lower threshold for admission. A further limitation of our study is that our cases were defined based on positive tests for SARS-CoV-2. Sensitivity of polymerase chain reaction tests for SARS-CoV-2 is 80%-90% depending on testing strategy.^25^ As such a proportion of true cases would have been misclassified. Finally, given that our healthcare worker population were predominantly white, our analysis lacked power to comment on the risk hospitalisation in ethnic minority groups.^26^

As the Northern Hemisphere enters winter and non-pharmacological measures on populations are relaxed, governments, healthcare managers and occupation health specialists need to consider how best to protect healthcare workers in the event of a resurgent pandemic. This is necessary to protect the healthcare worker and their families^13^ in addition to reducing onward transmission into the community.^4 12^ Our findings from the “first wave” in Scotland shows that healthcare workers in patient facing roles – especially those in “front-door” roles – are along with their households at particular risk. Crucially, those in non-patient facing roles had similar risks to that of the general population. These findings should inform decisions about the organisation of health services, the use of personal protective equipment and decisions about redeployment.

### Data sharing

Analysis code will be made available here – [WILL BE MADE PUBLIC PRIOR TO MANUSCRIPT PUBLICATION]. Since our analysis involved data on unconsented participants, we are unable to share individual level data.

### Transparency declaration

The corresponding author affirms that this manuscript is an honest, accurate, and transparent account of the study being reported; that no important aspects of the study have been omitted; and that any discrepancies from the study as planned have been explained.

### Summary boxes

#### Section 1: What is already known on this topic

Several systematic reviews and reports were identified summarising studies of COVID-19 infections in healthcare workers. The majority of studies related to COVID-19 infections in healthcare workers have been small, based in single centres, cross-sectional in nature, used methods highly susceptible to bias or restricted their populations to physicians and nurses. We found no studies evaluating the risk of COVID-19 infection on the risk to household members of healthcare workers.

#### Section 2: What does this study add

Healthcare workers and their households contribute a sixth of hospitalized COVID-19 cases. Our findings from the “first wave” in Scotland shows that healthcare workers in patient facing roles – especially those in “front-door” roles – are along with their households at particular risk of COVID-19 hospitalization. Crucially, those in non-patient facing roles had similar risks to that of the general population. These findings should inform decisions about the organisation of health services, the use of personal protective equipment and decisions about redeployment.

## Data Availability

Analysis code will be made available on an online repository on publication. Since our analysis involved data on unconsented participants, we are unable to share individual level data. This project was approved by the Public Benefit and Privacy Panel (2021-0013) in Scotland.

## Acknowledgements

Anoop Shah is funded via the British Heart Foundation through an Intermediate Clinical Research Fellowship (FS/19/17/34172) and David McAllister is funded via a Wellcome Trust Intermediate Clinical Fellowship and Beit Fellowship (201492/Z/16/Z). We would like to thank Colin Tilley, Peter Ward and Morag Macpherson provided access to the SWISS database, Kathy Kenmuir, Ben Hall Lucy Munro and Susie Dodds who provided advice on re-organisations in primary and secondary care, Frances Mair and Peter Hanlon who provided a front-line perspective on general practice and Kate Hughes who provided a front-line perspective on secondary care, and members of the Public Health Scotland COVID-19 Health Protection Study Group (Alice Whettlock, Allan McLeod, Andrew Gasiorowski, Andrew Merrick, Andy McAuley, April Went, Calum Purdie, Colin Ramsay, David Bailey, David Henderson, Diogo Marques, Eisin McDonald, Genna Drennan, Graeme Gowans, Graeme Reid, Heather Murdoch, Jade Carruthers, Janet Fleming, Jade Carruthers, Joseph Jasperse, Josie Murray, Karen Heatlie, Lindsay Mathie, Lorraine Donaldson, Martin Paton, Martin Reid, Melissa Llano, Michelle Murphy-Hall, Paul Smith, Ros Hall, Ross Cameron, Susan Brownlie, Adam Gaffney, Aynsley Milne, Christopher Sullivan, Edward McArdle, Elaine Glass, Johanna Young, William Malcolm and Jodie McCoubrey).

## References

1. European Centre for Disease Prevention and Control. COVID-19 situation update worldwide, as of 11 July 2020 2020 [Available from: https://www.ecdc.europa.eu/en/geographical-distribution-2019-ncov-cases accessed 12 July 2020.

2. Chou R, Dana T, Buckley DI, et al. Epidemiology of and Risk Factors for Coronavirus Infection in Health Care Workers. Ann Intern Med 2020 doi: 10.7326/M20-1632 [published Online First: 2020/05/06]

3. Wang D, Hu B, Hu C, et al. Clinical Characteristics of 138 Hospitalized Patients With 2019 Novel Coronavirus-Infected Pneumonia in Wuhan, China. JAMA 2020 doi: 10.1001/jama.2020.1585 [published Online First: 2020/02/08]

4. Adams JG, Walls RM. Supporting the Health Care Workforce During the COVID-19 Global Epidemic. JAMA 2020 doi: 10.1001/jama.2020.3972 [published Online First: 2020/03/13]

5. Nguyen LH, Drew DA, Joshi AD, et al. Risk of COVID-19 among frontline healthcare workers and the general community: a prospective cohort study. medRxiv 2020 doi: 10.1101/2020.04.29.20084111 [published Online First: 2020/06/09]

6. McAllister DA, Read SH, Kerssens J, et al. Incidence of Hospitalization for Heart Failure and Case-Fatality Among 3.25 Million People With and Without Diabetes Mellitus. Circulation 2018;138(24):2774–86. doi: 10.1161/CIRCULATIONAHA.118.034986 [published Online First: 2018/06/29]

7. Shah ASV, McAllister DA, Gallacher P, et al. Incidence, Microbiology, and Outcomes in Patients Hospitalized With Infective Endocarditis. Circulation 2020;141(25):2067–77. doi: 10.1161/CIRCULATIONAHA.119.044913 [published Online First: 2020/05/16]

8. Shah ASV, Anand A, Strachan FE, et al. High-sensitivity troponin in the evaluation of patients with suspected acute coronary syndrome: a stepped-wedge, cluster-randomised controlled trial. Lancet 2018;392(10151):919–28. doi: 10.1016/s0140-6736(18)31923-8 [published Online First: 2018/09/02]

9. McKeigue PM, Weir A, Bishop J, et al. Rapid Epidemiological Analysis of Comorbidities and Treatments as risk factors for COVID-19 in Scotland (REACT-SCOT): a population-based case-control study. medRxiv 2020 doi: https://doi.org/10.1101/2020.05.28.20115394

10. Scottish Government. Scottish Index of Multiple Deprivation Technical Notes. [Available from: https://www.gov.scot/collections/scottish-index-of-multiple-deprivation-2020/ accessed 25 July 2020.

11. Mateos P, Longley PA, O’Sullivan D. Ethnicity and population structure in personal naming networks. PLoS One 2011;6(9):e22943. doi: 10.1371/journal.pone.0022943 [published Online First: 2011/09/13]

12. McMichael TM, Currie DW, Clark S, et al. Epidemiology of Covid-19 in a Long-Term Care Facility in King County, Washington. N Engl J Med 2020;382(21):2005–11. doi: 10.1056/NEJMoa2005412 [published Online First: 2020/03/29]

13. McConnell D. Balancing the duty to treat with the duty to family in the context of the COVID-19 pandemic. J Med Ethics 2020;46(6):360–63. doi: 10.1136/medethics-2020-106250 [published Online First: 2020/04/26]

14. Rapid Response Report: Are healthcare workers at increased risk of COVID-19? Scientific Advisory Group Recommendation, Guidance, 2020. [Available from: https://www.albertahealthservices.ca/assets/info/ppih/if-ppih-covid-19-hcw-risk-rapid-review.pdf accessed 29th June 2020]

15. Pouwels KB, House T, Robotham JV, et al. Community prevalence of SARS-CoV-2 in England: Results from the ONS Coronavirus Infection Survey Pilot. doi: https://doi.org/10.1101/2020.07.06.20147348

16. Pollan M, Perez-Gomez B, Pastor-Barriuso R, et al. Prevalence of SARS-CoV-2 in Spain (ENE-COVID): a nationwide, population-based seroepidemiological study. Lancet 2020 doi: 10.1016/S0140-6736(20)31483-5 [published Online First: 2020/07/10]

17. Pan A, Liu L, Wang C, et al. Association of Public Health Interventions With the Epidemiology of the COVID-19 Outbreak in Wuhan, China. JAMA 2020 doi: 10.1001/jama.2020.6130 [published Online First: 2020/04/11]

18. Treibel TA, Manisty C, Burton M, et al. COVID-19: PCR screening of asymptomatic health-care workers at London hospital. Lancet 2020;395(10237):1608–10. doi: 10.1016/S0140-6736(20)31100-4 [published Online First: 2020/05/14]

19. Houlihan C, Vora N, Byrne T, et al. Pandemic peak SARS-CoV-2 infection and seroconversion rates in London frontline health-care workers. Lancet 2020 doi: https://doi.org/10.1016/S0140-6736(20)31484-7

20. Novel coronavirus (COVID-19) Guidance for secondary care: Management of possible/confirmed COVID-19 patients presenting to secondary care: Health Protection Scotland, 2020. [Available from: https://hpspubsrepo.blob.core.windows.net/hps-website/nss/2936/documents/1_covid-19-guidance-for-secondary-care.pdf accessed 28 th July 2020]

21. Liu M, Cheng SZ, Xu KW, et al. Use of personal protective equipment against coronavirus disease 2019 by healthcare professionals in Wuhan, China: cross sectional study. BMJ 2020;369:m2195. doi: 10.1136/bmj.m2195 [published Online First: 2020/06/12]

22. Avo C, Cawthorne KR, Walters J, et al. An Observational Study to Identify Types of Personal Protective Equipment Breaches on Inpatient Wards. J Hosp Infect 2020 doi: 10.1016/j.jhin.2020.06.024 [published Online First: 2020/06/27]

23. Houghton C, Meskell P, Delaney H, et al. Barriers and facilitators to healthcare workers’ adherence with infection prevention and control (IPC) guidelines for respiratory infectious diseases: a rapid qualitative evidence synthesis. Cochrane Database Syst Rev 2020;4:CD013582. doi: 10.1002/14651858.CD013582 [published Online First: 2020/04/22]

24. Coronavirus (COVID-19) related deaths by occupation, England and Wales: deaths registered up to and including 20 April 2020: Office for national statistics, 2020. [Available from https://www.ons.gov.uk/peoplepopulationandcommunity/healthandsocialcare/causesofdeath/bulletins/coronaviruscovid19relateddeathsbyoccupationenglandandwales/deathsregistereduptoandincluding20april2020 accessed 05th July 2020]

25. Williams TC, Wastnedge E, McAllister G, et al. Sensitivity of RT-PCR testing of upper respiratory tract samples for SARS-CoV-2 in hospitalised patients: a retrospective cohort study. 2020 doi: https://doi.org/10.1101/2020.06.19.20135756

26. Ethnicity and Outcomes from COVID-19: The ISARIC CCP-UK Prospective Observational Cohort Study of Hospitalised Patients. doi: http://dx.doi.org/10.2139/ssrn.3618215

